# The Costs and Benefits of Covid-19 Lockdowns in New Zealand

**DOI:** 10.1101/2021.07.15.21260606

**Authors:** Martin Lally

## Abstract

This paper considers the costs and benefits of New Zealand’s Covid-19 nation-wide lockdown strategy relative to pursuit of a mitigation strategy in March 2020. Using data available up to 28 June 2021, the estimated additional deaths from a mitigation strategy are 1,750 to 4,600, implying a Cost per Quality Adjusted Life Year saved by locking down in March 2020 of at least 13 times the generally employed threshold figure of $62,000 for health interventions in New Zealand; the lockdowns do not then seem to have been justified by reference to the standard benchmark. Using only data available to the New Zealand government in March 2020, the ratio is similar and therefore the same conclusion holds that the nation-wide lockdown strategy was not warranted. Looking forwards from 28 June 2021, if a new outbreak occurs that cannot be suppressed without a nation-wide lockdown, the death toll from adopting a mitigation strategy at this point would be even less than had it done so in March 2020, due to the vaccination campaign and because the period over which the virus would then inflict casualties would now be much less than the period from March 2020; this would favour a mitigation policy even more strongly than in March 2020. This approach of assessing the savings in quality adjusted life years and comparing them to a standard benchmark figure ensures that all quality adjusted life years saved by various health interventions are treated equally, which accords with the ethical principle of equity across people.

## 1. Introduction

As with most other countries, in early 2020 and in response to Covid-19, the New Zealand government implemented substantial general restrictions on mobility, with consequent significant curtailments of economic activities (“lockdowns”). Since then, the curtailments have been substantially relaxed, apart from being substantially but temporarily reinstated for the Auckland area. This paper attempts to assess the comparative merits of this lockdown strategy and a milder (“mitigation”) strategy involving case isolation, quarantining of members of their households, limiting large gatherings, social distancing, the wearing of masks on public transport, and restrictions targeted at only high-risk groups. The paper commences by examining this issue using data available as at 28 June 2021. It then considers the optimal choice based on information available at the time of the initial lockdown decision. Finally, it considers the optimal course of action if a new outbreak occurs that cannot be contained without resort to lockdowns.

## 2. The Costs and Benefits of Lockdowns in March 2020

### 2.1 Deaths

The primary purpose of lockdowns is to reduce deaths, and the deaths suffered under the lockdown policy are known. Much less clear is what the death toll would have been under a mitigation policy. In March 2020, Blakely et al (2020) estimated deaths from an eradication (lockdown) policy at 500, those from a mitigation strategy at 6,500 – 13,000, and those from no mitigation measures at 30,100 arising from 60% of the population then being infected. In November Binny et al (2020, Table 3) estimated the death toll from adoption of only Level 2 (a version of mitigation) at 32,000 (and more if the health system were saturated with cases, as it would be at that casualty level). Blakely et al’s (2020) estimates use mortality rates by age group from the Ferguson et al (2020) study in the UK. These in turn are based upon an epidemiological model in which each infected individual is estimated to infect *R* others (the reproduction rate) and this process extrapolated until so many people are infected that the virus dies out for lack of new targets. The estimates of Binny et al (2020) share this crucial feature.

In the first of these models, the estimated deaths under lockdown (500) are far in excess of the actual deaths incurred to date under lockdown, and this implies that their estimates of the additional deaths under mitigation will also be far too high. In addition, the death rate estimated by Binny et al (2020) under a mitigation policy (at least 32,000 deaths, which is 6,400 per 1m of New Zealand’s population of 5m) is vastly in excess of the death rate per 1m to date in any country pursuing a mitigation policy or any alternative.^1^ In addition, both of these models do not allow for the fact that, as the number of deaths rises, people will react by engaging in more and more protective actions that will reduce the future death rate, such as hand washing, mask wearing, reducing social interactions, working from home, etc.

In view of these problems, I estimate the additional deaths in New Zealand under a mitigation rather than a lockdown strategy by examining the death rates in other countries. Foster (2020) uses the death rate in Sweden to estimate the additional Australian deaths under a mitigation approach. However, Sweden was not the only mitigator; Iceland, Finland and Latvia did likewise. Even better would be to use the full set of countries with reliable data. One such approach would be to conduct a cross-country regression of death rates on variables found to influence such death rates, and include amongst the explanatory variables the strength of government restrictions. The coefficient on this latter variable would then provide an estimate of how many extra deaths would arise if the restrictions were less onerous. Chaudhry et al (2020) examines the 50 countries with the highest case counts as at 1 April 2020, and regresses cross-country death rates per 1m of population (up to 1 May 2020) on a number of independent variables, including various measures of government intervention, and find that *none* of these latter variables were statistically significant. Gibson (2020) conducts a similar analysis, using the 34 OECD member countries, death rates up to 18 August 2020, and various independent variables including the average level of government restrictions over the period of the crisis. He finds that policy stringency (averaged over the whole crisis period) is *not* statistically significant in explaining cross-country variation in death rates (ibid, Table 2), but average stringency prior to the estimated infection peak is mildly statistically significant along with a negative coefficient (ibid, Table 2). Hale et al (2020b) conduct a similar analysis, using 170 countries and data to 27 May 2020, and find that both the speed of government response (number of days from the first reported case till the government restrictions reach 40 on the Hale et al, 2020a Stringency index) and the severity of the restrictions (using the Stringency index of Hale et al, 2020a) reduce death rates.

Given the actual or probable unreliability of data from many countries, it is desirable to limit the analysis to countries for which the data is likely to be very reliable. It is also desirable to eliminate countries with federal systems, in which restrictions varied by state or province, because the Hale et al (2020a) data used to quantify restrictions is only available at the country-level. This leaves European countries and the East Asian democracies (Japan, South Korea, Taiwan, Singapore, plus Hong Kong). Hale et al (2020a) have constructed a set of indexes, which assign a daily score to each country for the severity of their restrictions imposed by government, ranging from 0 to 100 and taking account of different types of restrictions. I use their Stringency Index, which takes account of 8 different types of government restrictions.^2^ Death rates per 1m of population are drawn from https://www.worldometers.info/coronavirus/. All five East Asian countries have very low death rates regardless of the severity of restrictions, and the possible reasons (a culture of mask wearing, not shaking hands, compliance with government directives, extensive contact tracing and testing, and pre-existing immunity) are or were not applicable to the same degree in New Zealand. So, I use only the European countries, of which there are 33.^3^ They are similar (on average) to New Zealand in ethnicity, cultural norms, demographics, GDP per capita, and the quality of their health care systems.

In using the Stringency Index, there is a choice of the average and maximum values, and both have merits. The maximum reflects only government policy on one day whilst the average (crudely) takes account of it over the entire period of the crisis. However, a given average value could arise from a wide range of different policies. An extreme case of this would arise if one country adopted its maximum stringency index value of 100 on the first day, retained it for six weeks and then removed all restrictions because eradication had been achieved, whilst a second country maintained a stringency index value of 50 throughout the 12 weeks of the analysis. Both would have an average Stringency of 50, but would have adopted entirely different policies. I therefore use both the average and maximum values.

Death rates are likely to be affected by many variables other than the severity of government restrictions, and it is desirable to include them. I consider

a. population density (higher values increase the transmission rate of the virus),
b. the date of the first death (in days after the first recorded death on 15 February in France), because later dates provide more time for people, doctors and their governments to learn from others and adjust their behavior, ^4^
c. population (higher values provide a higher pool of virus targets before national borders constrict the movement of people and therefore the transmission of the virus),
d. GDP per capita (as a proxy for the quality of the health care system),
e. the population proportion over 65 (higher values imply a larger proportion of the population in the high risk group),
f. the average household size (higher values increase the pool of virus targets before household borders restrict interactions and therefore the transmission of the virus),
g. the number of nursing and elderly home beds per 100,000 of population (because higher values implies a higher concentration in the high-risk group, which increases or lowers the death rate depending upon the effectiveness of the quarantine procedures).
h. Flu intensity in the last two flu seasons.^5^

Mass vaccination commenced in Europe in December 2020. Since the speed of vaccination is likely to be a significant explanatory variable from that point, I conduct the analysis described above using data only to 30 December 2020. The first two variables are statistically significant, and substantially raise the adjusted *R*2, whilst the last five variables (added and tested separately) are not statistically significant and their inclusion each lowers the adjusted *R*^2^. I therefore retain only the first two variables. Regressing the death rate per 1m (*D*) up to 31 December on the maximum Stringency Index value (*S*), the population density (*PD*, in millions per 1,000 square miles), and date of first death (*FD*, in days from 15 February) yields the following result:

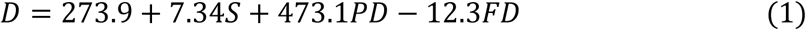

The *R*^2^ is 0.29, and the *p* values are 0.66, 0.27, 0.10 and 0.10 respectively. The coefficient on *S* is statistically insignificant and the sign on it is ‘wrong’ (positive rather than negative).^6^

This result may seem counterintuitive, but explanations are available. One possibility is that reverse causality applies, i.e., the choice of policy is influenced by the death rate as well as the death rate being affected by the policy choice. The Appendix investigates this possibility and concludes that it does not operate. A second possibility is that, even without government restrictions, people will take actions to lower their risks in a pandemic and the incremental effect of government actions (to the extent they are complied with) may then be too little to be statistically significant. A third possibility is that lockdowns induce some behaviours that *increase* the death rate, such as young people returning to live with their older parents, due to loss of their job or closure of the university they were attending, and if already infected to thereby infect their parents, who are at much greater risk of death. A fourth possibility is that some of the European lockdowns were not instituted quickly enough to be effective, and all of those that were instituted quickly enough were relaxed before eradication had been achieved (because their land borders were too porous to achieve eradication within an acceptable period) leading to a resurgence in cases when the lockdowns were relaxed.

This last possibility is very relevant here because New Zealand’s lockdown strategy succeeded in suppressing the two major outbreaks, leading to a very low death rate (of only 5 per 1m up to 30 December 2020). By contrast, European countries experienced

a. mitigation strategies (Finland, Iceland, Latvia, and Sweden, with death rates ranging from 82 to 861 per 1m up to 30 December), or
b. lockdown strategies that failed to suppress the virus (the rest, with death rates ranging from 80 to 1,656 per 1m up to 30 December).

Because the European data contains only cases of these two types, it does not provide an estimate of the death rate difference between a lockdown strategy that substantially succeeded and mitigation, and this differential is required for New Zealand. Accordingly, the coefficient on *S* in equation (1) is not useful for New Zealand. Accordingly, a different approach is required, as follows.

Since the evidence presented above is that lockdowns were not effective in Europe it follows that the European data is equivalent to that from a set of countries that pursued mitigation. So, an estimate of the New Zealand death rate under mitigation would be the average European death rate (680 per 1m as at 30 December), corrected for differences in variables that are statistically significant in explaining the death rate. For Europe, these are population density (*PD*) and date of first death (*FD*), and the resulting model using death rate data to 30 December is

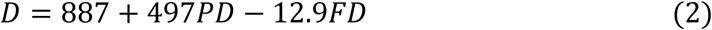

The *R*^2^ is 0.26 and the *p* values on the coefficients are 0.002, 0.09 and 0.09 respectively. Substitution of New Zealand’s values for the regressors, of *PD* = .0485 and *FD* = 42, yields an estimated death rate under mitigation of 369 per 1m.

This analysis uses data from European countries. The next best source of data appears to be that from the Americas, but with the exclusion of the US and Canada (because they are federal systems with variation in policy by state) and exclusion also of Cuba, Nicaragua and Venezuela (because authoritarian regimes are likely to deliberately understate deaths). I also exclude countries with less than 50,000 people because death rates expressed per 1m of population (as the data source does) are only then expressible in multiples of 20 or more. Across the countries for which both Hale et al (2020a) provides the Stringency data and the www.worldometers.info website provides death rate data, there are 28 countries: Brazil, Argentina, Colombia, Mexico, Peru, Chile, Ecuador, Bolivia, Panama, Dominican Republic, Costa Rica, Guatemala, Honduras, Paraguay, El Salvador, French Guyana, Jamaica, Haiti, Trinidad and Tobago, Suriname, Aruba, Guyana, Belize, Uruguay, Cayman Islands, Barbados, Bermuda, and Dominica. Unlike the European data, population density and date of first death are not statistically significant, but the following two regressors are:

a. population (higher values provide a higher pool of virus targets before national borders constrict the movement of people and therefore the transmission of the virus),
b. having no land borders with other countries (water barriers rather than land borders better restrict the flow of people and hence the virus into a country).

If the maximum Stringency index is added, it is not statistically significant and the estimated coefficient on it is positive rather than the expected negative. With *P* denoting population in millions and *I* denoting no land borders (1 if so and 0 otherwise), the model exclusive of *S* is thus:

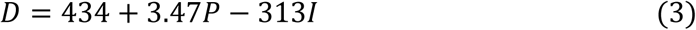

The *R*^2^ is a respectable 0.43, and the *p* values on the three coefficients are 0, 0.01 and 0.01 respectively. Substitution of New Zealand’s values for the regressors, of *P* = 5 and *I* = 1, yields an estimated death rate under mitigation of 138 per 1m.

If the European and Americas data are pooled, *P*, *I* and date of first death (*FD*) are statistically significant at the 10% level, with all coefficients having the expected signs. Addition of *S* yields a coefficient that is not statistically significant, and with a positive coefficient. The resulting model exclusive of *S* is thus:

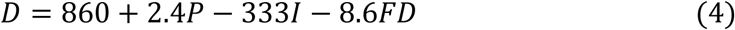

The *R*^2^ is a respectable 0.36, and the *p* values on the four coefficients are 0, 0.07, 0.02 and 0.02 respectively. Substitution of New Zealand’s values for the regressors, of *P* = 5, *I* = 1, and *FD* = 42, yields an estimated death rate under mitigation of 178 per 1m.

Across these three models (2), (3) and (4), the estimated death rate for New Zealand under a mitigation policy is 138 to 369 per 1m. With 5m people in New Zealand, this implies 700 – 1,850 deaths in New Zealand up to 30 December if a mitigation policy had been pursued. The estimate of 1,850 uses the best quality data (from Europe) but is likely to be too high because New Zealand is an island group, and this reduces its death rate, but there are too few islands in the European data (only two) for the dummy variable “Island” to be statistically significant and therefore warrant inclusion in equation (2). By comparison, the average death toll amongst the four European mitigators was 340 per 1m, their average values for the independent variables in equation (2) is less favourable than for New Zealand, and only one of them is an island; this further supports the conclusion that the death toll for New Zealand under a mitigation policy would have been no more than 369 per 1m to 30 December.

These estimates presume that Covid-19 deaths are accurately recorded. However, some Covid-19 deaths may be mistakenly attributed to other causes, or deaths from other causes mistakenly attributed to Covid-19, with the latter error possible simply because most victims have co-morbidities. In addition, lockdown discourages or prevents some people suffering from non Covid-19 conditions from seeking medical attention, leading to some deaths in lockdown countries that would not otherwise have occurred, and these should be included in the incremental deaths from lockdown. In addition, mitigation increases the load on hospitals, leading to more deaths from other causes (through lack of care) in mitigation countries, and these should be included in the incremental deaths from mitigation. An estimate of the Covid-19 deaths that accounts for all of these phenomena is the actual deaths in 2020 less the predicted number sans Covid-19 (“Excess Deaths”). The Euromomo Network has done so and estimated the number of deaths across 18 European countries progressively through 2020, 2019 and 2018 relative to a prediction (“baseline”). The Excess Deaths for 2020 exhibit sharp increases in March-April and November-December, consistent with the pandemic. The Excess Deaths to 31 December relative to the baseline are 290,000 for 2020 (from 15 February when the first Covid-19 death occurred in any of these 18 countries), 70,000 for 2019 and 115,000 for 2018.^7^ By contrast, the deaths attributed to Covid-19 across these 18 countries (to 31 December) were 334,000. Thus, if the baseline were used, the Excess Deaths in 2020 would be 290,000 and therefore the deaths attributed to Covid-19 of 334,000 would be too high by 15%. However, the baseline is an imperfect prediction, as evidenced by the results for 2018 and 2019 (which would be zero if the predictions were accurate). This is simply a consequence of the fact that deaths in these countries in a typical year are about 3m, so that the prediction error of 115,000 for 2018 is a small proportion.^8^ All of this suggests that the deaths attributed to Covid-19 are approximately correct.

Turning now to further deaths from 30 December 2020, these are likely to be significantly influenced by the speed of vaccination. However, even if a model could be developed to quantify this, the speed with which New Zealand would have vaccinated had it followed a mitigation strategy is indeterminable (and likely to be much faster than the speed with which it is vaccinating under its lockdown policy). So, the best available estimate of the additional deaths in New Zealand from 30 December 2020 under a mitigation strategy would seem to be that arising from applying the average ratio for Europe. Over the period from 30 December 2020 to 28 June 2021, the European deaths are 95% of those to 30 December, and the rate of deaths fell sharply towards the end of the period. This suggests that deaths from 30 December 2020 until the end of 2021 (at which point mass vaccination is likely to have been largely completed) will not exceed 1.5 times those to 30 December 2020.^9^

Applying this ratio of 1.5 to New Zealand, with estimated deaths to 30 December 2020 under a mitigation strategy of 700 – 1,850, yields 1,750 – 4,600 deaths to the end of 2021. By contrast, deaths to 28 June 2021 under a lockdown policy have been 26. So, the extra deaths resulting from a mitigation policy rather than a lockdown policy are estimated to be 1,750 – 4,600. By contrast, Blakely et al (2020) predicted an extra 6,000 – 12,500.

### 2.2 Quality Adjusted Life Years

In assessing the merits of health interventions, the standard methodology amongst health economists is to multiply the expected lives saved from a health intervention by the average residual life span of the victims sans intervention, to yield the “Life Years” saved by the intervention, followed by some discount if the quality of these life years saved would be less than that of a normal healthy person. The result is called the Quality Adjusted Life Years (QALY) saved by the intervention, which is then compared to a benchmark figure.

Blakely et al (2020) adopt an average residual life expectancy for the victims of five years, but provide no supporting evidence. Heatley (2020, page 5) estimates the average age of Covid-19 victims in Italy at 79.5 years, notes that a New Zealander of that age has a residual life expectancy of 10.7 years, and that allowance for the greater incidence of co-morbidities warrants a reduction in their residual life expectancy to five years.

More detailed analysis of European data supports this figure of five years. I illustrate this with Sweden, which adopted a mitigation policy and incurred the highest death rate amongst countries that did so. The age distribution of the Covid-19 victims is shown in the first two columns of Table 1, and the residual life expectancy (RLE) of Swedish people in each such age group is shown in the third column. Using this data, the average residual life expectancy of Swedish people with the same age distribution as the Covid-19 victims is 10.9 years.^10^ However, the Covid-19 victims differ from Swedish people of the same age distribution in two very significant ways.

**Table 1:** Residual Life Expectancy of Swedish Covid-19 Victims

| Age Group | Victims | RLE | RLE | RLE | RLE |
| --- | --- | --- | --- | --- | --- |
| 0-9 | 2 | 79.55 | 79.55 | 79.55(0.7) | 79.55(0.7) |
| 20-29 | 10 | 60.25 | 60.25 | 60.25(0.7) | 60.35(0.7) |
| 30-39 | 19 | 50.5 | 50.5 | 50.5(0.7) | 50.5(0.7) |
| 40-49 | 45 | 40.8 | 40.8 | 40.8(0.7) | 40.8(0.7) |
| 50-59 | 164 | 31.3 | 31.3 | 31.3(0.7) | 31.3(0.7) |
| 60-69 | 406 | 22.4 | 22.4 | 22.4(0.7) | 22.4(0.7) |
| 70-79 | 1268 (22%) | 14.3 | 14.3 | 14.3(0.7) | 4.45 |
| 80-84 | 1219 (21%) | 9.1 | 6.67 | 4.72 | 2.80 |
| 85+ | 2747 (47%) | 6.3 | 0.6 | 0.6 | 2.14 |
| Average (yrs) |  | 10.9 | 7.7 | 5.5 | 4.7 |

The first of these differences is that a large proportion of the victims were residents of nursing homes, whose average residual life expectancy sans Covid-19 was very low and might be even lower than suggested by their ages. If so, conditioning on residency of a nursing home as well as age would reduce the average residual life expectancy of the victims. Stern and Klein (2020, page 5) estimate that 53% of the Swedish Covid-19 victims aged at least 70 were residents of nursing homes, and that their average residual life span sans Covid-19 was only seven months (ibid, pp. 16-17). Conservatively treating this subset of victims as the oldest in Table 1, they represent the entire 85+ group (47%) plus additional victims in the 80-84 group constituting 6% of the entire set of victims (6/21 of that group). Replacing the residual life expectancy of these people by seven months (0.6 years), the average residual life expectancy calculated from the data in Table 1 would fall to 7.7 years as shown in the fourth column of Table 1.^11^ By contrast, if this nursing home group were spread through the 70+ groups in proportion to the size of these groups, 28% would be in the 85+ group, 13% in the 80-84 group, and 13% in the 70-79 group. Replacing the residual life expectancy of these people by 0.6 years, the average residual life expectancy calculated from the data in Table 1 would fall to 6.4 years.

The second unusual feature of these Covid-19 victims is that they were unusually unwell, even for their age; virtually all had at least one co-morbidity, which is presumably well in excess of the rate for the general population of the same age/sex distribution.^12^ Accordingly, a lower bound on the impact of this factor is the increase in the mortality risk of a person suffering from one of these co-morbidities relative to the general population of the same age and sex. A common such co-morbidity was type 2 diabetes. The British National Health Service provides estimates for the increase in mortality risk from this disease (relative to the general population) by age and sex (NHS, 2018, Figure 8). Averaging over these categories, the increase is about 50%. However the group of interest here excludes those in nursing homes, because the estimate for the residual life expectancy of these victims already reflects co-morbidities. This exclusion lowers the average age of the remaining victims, and suggests an increase in their mortality risk of about 80%. In addition, a person with a residual life expectancy of 10 years (the average for those of the same age as the Covid-19 victims) would have a current mortality risk of about 5% over the next year, growing at about 11% per year compounded:^13^

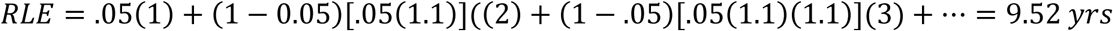

Raising this initial mortality risk by 80%, from 5% to 9%, along with the same growth rate of 11%, reduces the residual life expectancy from 9.52 yrs to 6.68 yrs, i.e., a reduction of 30%. A similar percentage reduction applies to the average residual life expectancy of a group. As noted above, virtually all of the victims had at least one co-morbidity, and multiple co-morbidities would reduce the average residual life expectancy of a group by even more than estimated here.

So, the subset of Swedish victims from nursing homes have their residual life expectancy set as before at 0.6 years (which will also reflect their co-morbidities), and all others have their residual life expectancy reduced by (conservatively) 30%. The results are shown in the penultimate column of Table 1, with the nursing home group conservatively assumed (as before) to be the oldest, and the last column of Table 1, in which the nursing home group is spread throughout the 70+ groups in proportion to their sizes.^14^ The average residual life spans are 5.5 and 4.7 years respectively, and a good estimate would lie between these figures. This supports the estimate of five years.

The last step here is the discount to reflect the imperfect health of virtually all of these victims sans Covid-19. Miles et al (2020, page 69) use 20% based on prevailing discounts for type 2 diabetes with and without additional problems. In particular, they cite Beaudet et al (2014, Table 3), who favour a quality of life discount of 21% for Type 2 diabetes without complications, and substantial additional discounts for further problems including 9% for heart disease and 16% for stroke. These discounts in Beaudet et al (2014) suggest that Miles et al’s (2020) 20% discount for an average covid-19 victim is low. Consistent with this, Briggs (2020, Figure 3) uses a discount of about 30% for Covid-19 victims, based upon norms arising from survey data from Szende et al (2014). Furthermore, a large proportion of the victims were residents of nursing homes, for which the quality of life discount could reasonably be even higher. I adopt a very conservative estimate of the discount, of 20%.

In conclusion, the QALYs saved by the New Zealand government pursuing lockdown rather than mitigation are estimated at up to 4,600*5*0.8 = 18,400. This estimate is likely to be too high, because the 4,600 additional deaths are likely to be too high and the 20% discount too low.

### 2.3 Expected GDP Losses

Turning now to the costs of the lockdown policy, this principally takes the form of lost GDP. Shortly before the pandemic arose, in December 2019, The Treasury (2019, page 3) forecasted New Zealand’s real GDP growth rates for 2020-2024 at the rates shown in the first row of Table 2.^15^ This is an estimate of growth in the absence of the pandemic. Arbitrarily designating 2019 GDP as 100, the GDP results under this path are shown in the next row of the table. In May 2021 they released updates as shown in the third row of the table (The Treasury, 2021, Table 1.1), with the implied GDP path in the fourth row. The last row of the table shows the difference between the two paths, which aggregates to 13.8, i.e., 13.8% of New Zealand’s 2019 GDP. Since New Zealand’s 2019 GDP was $311b, this is $43b.^16^ This estimate would seem to embody the full effect because the two real GDP forecast paths in Table 2 have fully converged over the period for which the forecasts are available (out to 2025). By comparison, Pujol (2020, Table 1) reports estimates of this type from nine advanced economies that locked down, and the median loss is 25%. In addition, Gomez-Pineda (2020, Figure 1) graphically presents annual estimates of this type for both advanced and developing economies (each averaged), and the accumulated losses over the 2020-24 period are 25% and 40% respectively.

**Table 2:**
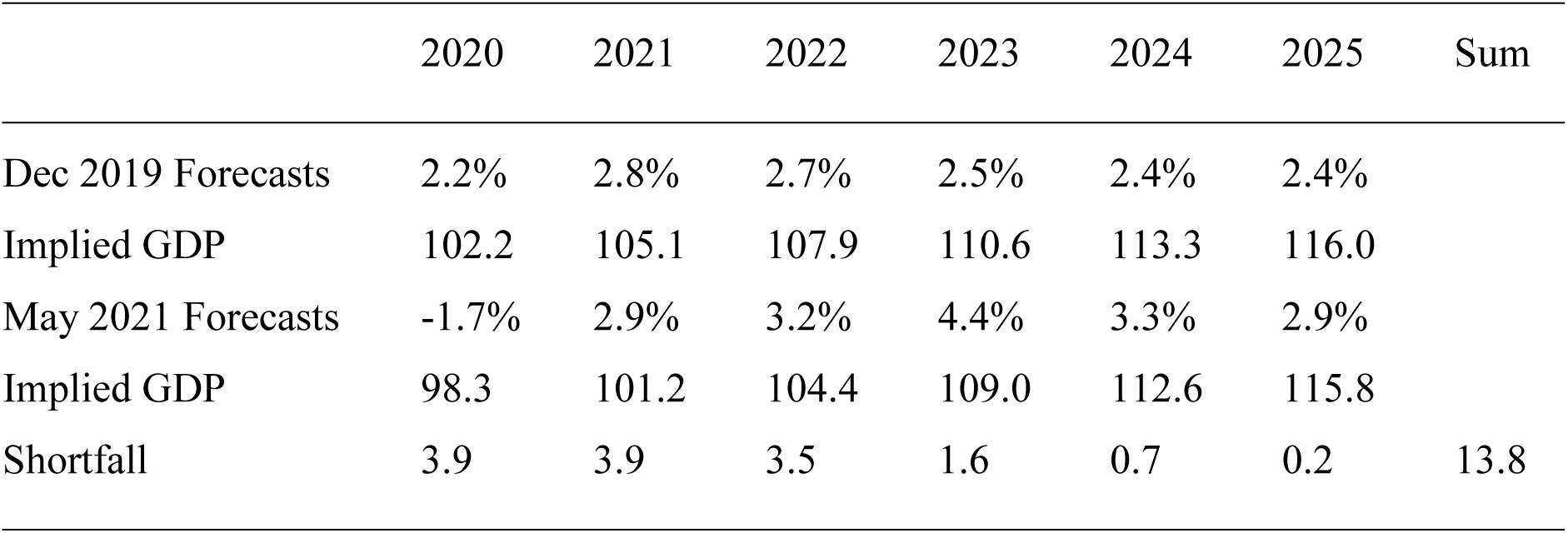
GDP Forecasts

|  | 2020 | 2021 | 2022 | 2023 | 2024 | 2025 | Sum |
| --- | --- | --- | --- | --- | --- | --- | --- |
| Dec 2019 Forecasts | 2.2% | 2.8% | 2.7% | 2.5% | 2.4% | 2.4% |  |
| Implied GDP | 102.2 | 105.1 | 107.9 | 110.6 | 113.3 | 116.0 |  |
| May 2021 Forecasts | -1.7% | 2.9% | 3.2% | 4.4% | 3.3% | 2.9% |  |
| Implied GDP | 98.3 | 101.2 | 104.4 | 109.0 | 112.6 | 115.8 |  |
| Shortfall | 3.9 | 3.9 | 3.5 | 1.6 | 0.7 | 0.2 | 13.8 |

Some of these GDP losses of $43b would have arisen without any New Zealand government-imposed lockdowns, because some people would have reduced their interactions with others anyway; for example, a foreigner electing not to make a trip to New Zealand that they would otherwise have made, or New Zealanders choosing to avoid cafes. Further losses would have arisen due to the additional actions of foreign governments, such as these governments preventing or discouraging their citizens from making foreign trips. Further losses would have arisen if the New Zealand government had followed merely a mitigation strategy, which includes border closures. Finally, further losses would have arisen from the New Zealand government instead following a lockdown strategy. It is only the last category of these losses that can be attributed to the New Zealand government choosing a lockdown rather than a mitigation policy.

Estimating the proportion arising from this last category is difficult. Andersen et al (2020) examine the drop in consumer spending in the early stages of the pandemic in both Denmark (which adopted a lockdown policy) and Sweden (which adopted a mitigation policy). They find that the drop in Sweden was 86% of that in Denmark (25% drop versus 29% drop), implying that 14% of the drop in Denmark was due to a lockdown rather than a mitigation policy. In a similar study (Goolsbee and Syverson, 2020) examine adjoining US counties with one area subject to lockdown and the other not; the drop in consumer activity in the latter area was 88% of the former, implying that only 12% was due to the lockdown. Doti (2021, page 120) conducts a cross-sectional study of all US states and concludes that 75% of the GDP losses were due to state government level interventions (from a Stringency Index of zero to the state average of 42), i.e., 2.2% out of 3%. Since the latter variation in the Stringency Index values approximates the difference between mitigation and extreme rather than an average lockdown, this suggests that an average lockdown would explain 1.1%/1.9% = 57% of the GDP loss. Aum et al (2020) estimate the effect of increased infections upon the unemployment rate in Korea (which did not lockdown), the US and UK (which did), and conclude that the effect is twice as great in the US and UK, leading to the conclusion that lockdowns explain 50% of the loss of employment. In a much broader study, the IMF (2020, Chapter 2) examined 28 countries and concluded that lockdowns contributed 40% of the reduction in ‘Google Mobility Data’ in advanced economies, which is a proxy for the GDP loss. In a similar study, Caselli et al (2021) found that the reduction was 44% using the same data source (ibid, Figure 4) and also 44% using job postings data (ibid, Figure 5), all for advanced economies. In an approach directly comparable with Table 2 above, Pujol (2020, Table 2) presents estimates of the GDP losses for nine advanced economies that locked down (US, Canada and seven Western European economies), and two that did not (Sweden and Japan); the median of the first group is 25% and that of the latter is 16%, which implies that 36% of the loss is due to lockdowns.

In summary, these estimates of the proportion of GDP losses due to an average lockdown relative to mitigation range from 12% to 57%. The best estimates here are the last three (40%, 36% and 44%), because they each cover a wide range of countries. I adopt the median estimate of 40%. Applying it to the New Zealand GDP loss of $43b yields a loss due to the lockdowns of $17b.

### 2.4 Cost per QALY Saved

In summary, the QALYs saved by locking down rather than mitigating are estimated at up to 18,400 whilst the associated GDP losses are expected to be $17b. If these GDP losses were the only cost of lockdowns, the cost per QALY saved would then at least $924,000 as follows:

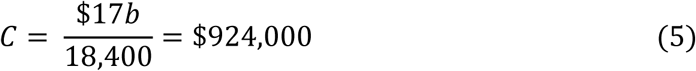

I now attempt to quantify additional costs of mitigation. Firstly, there are the medical costs of those requiring short-term hospitalization under a mitigation policy. Wilson (2020) estimates hospitalized cases at 146,000 and ICU cases as 36,600 under a “worst-case scenario” of 27,600 dead. Since my upper bound on the number of dead under a mitigation policy is 4,600, this implies 24,000 hospitalised cases and 6,000 ICU cases. I assume (to be conservative in respect of costs) that the New Zealand hospital system could accommodate all of them. Furthermore, whatever number of cases were accommodated, they would be to some degree permanently displacing other types of patients so the incremental costs would be even less.^17^ Even without displacement, most of these costs would be fixed (staff, buildings, and equipment) and therefore irrelevant. In the interests of being conservative, I assume no displacement of other types of patients and all costs being variable. Gros (2020, section 2.2) estimates the cost per patient for those requiring general hospital care at 30% of GDP per capita, and those requiring Intensive Care treatment at a further cost per patient of 60% of GDP per capita. With New Zealand’s GDP per capita of $62,000, the upper bound on the resulting incremental costs would be $0.7b as follows:

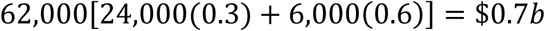

Substituted into equation (5) yields a cost per QALY of at least $890,000 as follows:

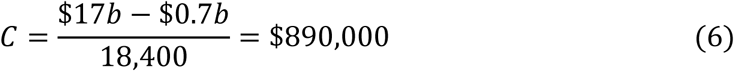

Secondly, mitigation gives rise to some survivors who may experience significant long-term adverse consequences. Using data from the Covid Symptom Study, Sudre et al (2020) estimate that 13.3% of those who tested positive remained unwell for at least four weeks, with 8.8% resolved in 4 - 8 weeks, a further 2.2% resolved in 8 – 12 weeks, and the remaining 2.3% unresolved after 12 weeks. A pattern consistent with this data is that, amongst this group who are still unwell after 4 weeks, 68% experience symptoms for 4 – 8 weeks, 16% for 8 – 12 weeks, 8% for 12 – 16 weeks, etc. The average time unwell is then

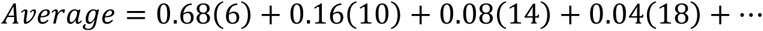

This series can be decomposed into a set of geometric progressions and then added, to yield an average of 9 weeks (0.16 years). As of 15 October 2020, matching the date of Sudre et al’s (2020) paper, there were 38.6m recorded cases and 1.1m deaths worldwide.^18^ So, the ratio of these long-recovery covid cases to deaths is 38.6m*0.133/1.1m = 4.7. Furthermore, consistent with the figure used earlier for Covid-19 victims suffering from serious pre-existing conditions, their quality of life is thereby reduced by 20%. Allowing for all this is then equivalent to increasing the QALYs saved from lockdown rather than mitigation by 4% as follows:

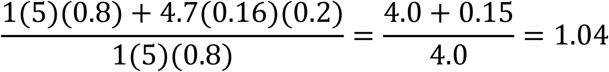

This raises the denominator in equation (6) by a factor of 1.04. There will also be medical costs associated with these longer term sufferers. For example, if each such person’s medical costs average $10,000 per year in which symptoms are experienced, the cost for 22,000 survivors (4,600 victims*4.7) for 0.16 years on average is $35m. Both the denominator adjustment of 1.04 and the numerator adjustment of $35m are too small to warrant inclusion in equation (6), and alternative (reasonable) assumptions about the time profile of the resolution of these cases and the cost per year per patient do not change this conclusion.

Thirdly, mitigation gives rise to work absences amongst those who are infected and must self-isolate. Gros (2020, section 2.1) assumes all members of a population are infected, 30% require a work absence of four weeks and a further 20% require six weeks, leading to a GDP loss of 5% of one year’s GDP:

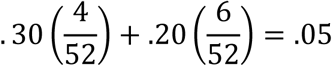

However the assumption that everyone in a population would become infected is excessive. Blakely et al (2020) estimate that the infection rate would not exceed 60% because the epidemic would by then peter out through herd immunity, Boyd (2020, page 3) adopts a base case of 40% based upon past pandemics, and Aguas et al (2020) estimates an even lower rate. Furthermore, even if Gros’s estimate of 50% of those infected requiring a work absence were correct, this does not yield a proportionate decline in GDP for various reasons. In particular, many of the people required to isolate could still perform their work from home. Furthermore, even where those isolated could not thereby perform their tasks for this period of weeks, other employees of the organization would increase their productivity or hours of work to at least partly compensate, and/or customers of the businesses would experience longer wait times with no loss of output, and/or the absent employees would be able to perform at least some of the work upon their return in addition to their normal workloads. Accordingly, a more reasonable estimate of the GDP loss than Gros’s would involve 40% of the population being infected, and 30% of those requiring isolation able to still perform their jobs at home, and 75% of the rest having their work performed by others or by them upon their return to work or addressed through longer customer queues. The resulting GDP loss would then be only 5%*0.4*0.7*0.25 = 0.35% rather than 5% of one year’s GDP. In dollar terms this is $311b*0.0035 = $1.1b. Modifying equation (6), the cost per QALY saved would be at least $826,000 as follows:

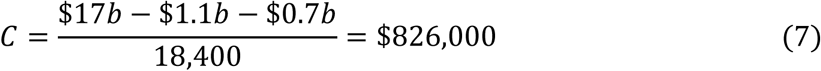

This cost per QALY is too low for two principal reasons. Firstly, the estimate of 18,400 QALYs saved by locking down is likely to be too high because it uses the highest estimate of additional deaths from locking down (from the European data). Secondly, no allowance has been made for various phenomena that would raise the costs of lockdowns but cannot readily be quantified: problems arising from the increased unemployment (addiction, crime, domestic violence, mental health problems, and premature death), loss of social interactions, increased anxiety, disruption to the education of the Covid-19 student cohort, and the deprival of liberties that people would otherwise enjoy. Lockdowns also disrupt the normal operation of the health care system, leading to deaths that would not otherwise occur (such as people who fail to have cancer screening tests done), but failure to lockdown may also saturate the system with covid-19 cases, leading to deaths amongst other types of patients who have been crowded out. The net effect of this point is unclear, because it depends inter alia on how quickly a country could expand its health care system to accommodate the increased caseload, but the net effect over the last ten months has been small because the deaths attributed to covid-19 approximate the excess deaths relative to pre-pandemic forecasts (as discussed in section 2.1).

Foster (2020) attempts to quantify the adverse psychological impact of lockdowns on the average Australian, and concludes that it dominates all other considerations. Such estimates are inherently subjective. Nevertheless, one aspect of this issue can be reasonably objectively assessed: the psychological effect of unemployment on the unemployed during the period of their unemployment. Table 3 shows forecast growth rates for the Labour Force in December 2019 (The Treasury, 2019, Table 1.1) and in May 2021 (The Treasury, 2021, Table 1.1).^19^ Arbitrarily designating the 2018-19 Labour Force as 100, the Labour Force results are shown under each forecast path, and the shortfall for each year shown in the last row, which aggregates to 6.3, i.e., 6.3% of New Zealand’s 2019 Labour Force. Since New Zealand’s 2019 Labour Force was 2.5m, this is the equivalent of 158,000 people unemployed for one year.^20^ This estimate is conservative because the two Labour Force forecast paths in Table 3 have not fully converged over the period for which the forecasts are available.

**Table 3:** Employment Forecasts

|  | 2020 | 2021 | 2022 | 2023 | 2024 | 2025 | Sum |
| --- | --- | --- | --- | --- | --- | --- | --- |
| Dec 2019 Forecasts | 1.4% | 2.1% | 1.9% | 1.6% | 1.5% | 1.5% |  |
| Implied Employment | 101.4 | 103.5 | 105.5 | 107.2 | 108.8 | 110.4 |  |
| May 2021 Forecasts | 1.7% | 0.2% | 1.4% | 2.4% | 1.9% | 1.8% |  |
| Implied Employment | 101.7 | 101.9 | 103.3 | 105.8 | 107.8 | 109.8 |  |
| Shortfall | -0.3 | 1.6 | 2.2 | 1.4 | 1.0 | 0.6 | 6.3 |

These Labour Force shortfalls arise from the pandemic, and it is only the fraction due to the lockdowns that are of interest. Section 2.3 estimate the proportion of the GDP shortfalls due to lockdowns at 40%, and the same proportion is applied here. So, lockdowns are expected to have reduced the size of the Labour Force by the equivalent of 158,000*0.4 = 63,000 for one year, i.e., 63,000 people lost their jobs for one year. Frijters (2020) estimates that the loss of employment for one year reduces a person’s quality of life during that year by the equivalent of 0.12 years of life.^21^ This estimate is subjective, but so too is the estimate of the reduction in life quality of a typical covid-19 victim by 20% to reflect their co-morbidities (in section 2.2 above). In the interests of being conservative, I halve the QALY impact of unemployment for one year from 0.12 years of life to 0.06 years of life. The loss of employment for 63,000 people for one year would then be equivalent to the loss of 63,000*0.06 = 3,800 QALYs. Netting this off against the 18,400 QALYs saved by the lockdown, and modifying equation (7), the cost per QALY saved by lockdown would then be at least $1.04m as follows:

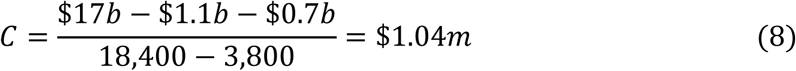

### 2.5 The Benchmark Valuation of a QALY

Turning now to the value of a QALY saved, Blakely et al (2020) used a QALY value of $45,000, consistent with the *BODE*^3^ project (which estimates the cost effectiveness of various health interventions in New Zealand). Kvizhinadze et al (2015, page 3) used the same figure for New Zealand, based upon a WHO recommendation of per capita GDP. However, with a current population of 5.0m and 2019 GDP of $311b, New Zealand’s current GDP per capita is $62,000. Pharmac (2015, page 12) does not prescribe a value, because other factors influence its decisions on subsidising drugs and any value would depend upon its budget. However, Pharmac’s highest average cost per QALY saved in recent years is $27,027 in 2016/17, which implies $33,306 in 2020.^22^ In evaluating the merits of extending the first Level 4 lockdown for five extra days, Heatley (2020, page 9) used the latter figure. In the interests of being conservative, I use the highest number here of $62,000.^23^

A related concept is the “Value of a Statistical Life” (VOSL), which values all lost years of an average aged person’s life. This is generally employed in decisions on road safety and the current value is $4.37m (Ministry of Transport, 2019, page iii). To convert to a value per year (called the Value of a Statistical Life Year or VOSLY), which can be compared to the value of a QALY, it would be necessary to first determine the average residual life span of a road accident victim. The median age of a victim is 41, and 2/3 are male.^24^ The median residual life span of a 41 year old is 43 years for a female and 40 for a male.^25^ This implies an average residual life span for a median aged road victim (of age 41) of 41 years. Applying Pharmac’s recommended discount rate of 1.58%, the value per QALY would be *V* such that^26^

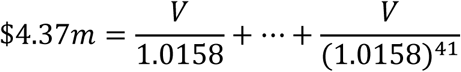

The solution is *V* = $146,000. This $4.37m estimate of the VOSL was derived from a 1991 survey (Ministry of Transport, 2019, page 14), as is usually the case for such estimates, whilst the value of a QALY comes from public health experts or deduced from the health expenditure decisions by government; the latter would then seem to be more relevant to the present situation involving a health policy choice by government. Consistent with this, Gros (2020, page 6) notes that it is the approach that is “practiced routinely by the medical profession” and Miles at al (2020, page 76) that it is consistent with the approach to other health expenditures. I therefore favour a QALY value of $62,000.

Interestingly, some analyses of the Covid-19 issue have been conducted by coupling the VOSL with the expected number of lives saved rather than coupling the value of a QALY or a VOSLY with the expected number of (quality adjusted) life years saved (for example, Chapple, 2020; Thunstrom, 2020; Holden and Preston, 2020). This may reflect a belief that Covid-19 victims have a typical average residual life span and perfect health sans Covid-19, which may be true in safety interventions but is not the case here, and would therefore significantly overestimate the benefits in QALYs saved. Alternatively, it may reflect their ethical belief that all lives saved are equally valuable, which implies that one would spend as much to extend the life of a person by one day (or even one hour) as one would spend to extend the life of a different person for fifty years. If the latter interpretation is correct, it would be inconsistent with prevailing views amongst public health experts in New Zealand and elsewhere, in which the impact of health interventions on the residual life expectancy of the targets is estimated and converted to a monetary figure using a value per year (Kvizhinadze et al, 2015; Bertram et al, 2016; Blakely et al, 2020).

### 2.6 Comparison of Cost with Benchmark

In summary, the cost per QALY saved is at least $826,000 if the psychological effects of unemployment are ignored and at least $1.04m if these latter effects are recognized, as shown in equations (7) and (8). Since the benchmark figure is $62,000 then the cost per QALY saved significantly exceeds this benchmark figure of $62,000, and therefore the March 2020 lockdowns were not justified. This conclusion is strengthened by numerous additional considerations. Firstly, using the VOSLY benchmark of $146,000 rather than the standard benchmark of $62,000 does not change the conclusion. Secondly, substituting Blakely et al’s (2020) higher estimate of the additional deaths from mitigation (up to 12,500) for the additional 4,600 deaths underlying equation (7) does not change the conclusion. Thirdly, many of the parameter estimates in equations (7) and (8) are towards the end of their probability distributions that yield the lowest possible cost per QALY, most particularly the 18,400 extra QALYs lost from mitigation (likely to be too high). Fourthly, apart from the allowance within equation (8) for the contemporaneous effect on the unemployed from losing their jobs, no allowance has been made for various phenomena that would raise the costs of lockdowns but cannot readily be quantified: more tangible problems arising from increased unemployment (addiction, crime, domestic violence, mental health problems, and premature death), the loss of social interactions, increased anxiety, disruption to the education of the Covid-19 student cohort, and the deprival of liberties that people would otherwise enjoy.

The conclusion here might seem to result from the mechanistic application of an economic rule, devoid of ethical considerations. However, consistent application of this rule is necessary to ensure that all quality adjusted life years are treated equally, which in turn derives from the ethical principle of equal treatment of people (appropriately adjusted for differences in their residual life expectancy and the quality of those years).

## 3. The Merits of Lockdown Versus Mitigation Using Data Available in March 2020

I now consider the merits of the lockdown decision in March 2020 using data available at that time. To be conservative, I focus upon equation (7). The denominator there must be replaced by an estimate derived from contemporaneous data. In March 2020, Blakely et al (2020) estimated the death toll under lockdown at 500 if successful, that under mitigation at 6,500 – 13,000, and 30,100 dead if no mitigating actions were taken arising from an infection rate of 60% of the population. Contemporaneously, James et al (2020, Table 2) presented predictions of the death tolls in New Zealand from a range of possible control strategies. Lockdown corresponds to the last strategy in their Table 2, and involves predicted deaths of 20 (0.0004% of the population). However, none of the other strategies examined by them in their Table 2 corresponds to “mitigation” as defined by Ferguson et al (2020) and Blakely et al (2020). The fourth strategy in their Table 2 embodies case isolation and quarantining of members of their households, but not also social distancing, the wearing of masks on public transport, and restrictions targeted at only high-risk groups; it therefore (unsurprisingly) yields a predicted death toll much higher than that of Blakely et al (2020), at 62,500. Table 2 also includes a no control strategy, with a predicted death toll of 83,000 (1.67% of the population). James et al (2020, page 7) do consider a strategy they describe as mitigation, with a predicted death toll of 25,000 (0.508% of the population), but it involves a combination of periods of low control (case isolation plus household quarantining) with periods of lockdown as required to keep the number of cases within the capacity of the hospital system. I therefore treat the work of Blakely et al (2020) as providing the best estimate of the death toll from mitigation that was available in March. So the best estimate in March 2020 of the death toll under mitigation was 9,750 (the midpoint of 6,500 and 13,000) and that under lockdown was 500. Assuming lockdown was bound to be successful, this implies (9,750 – 500)*5*0.8 = 37,000 QALYs saved by locking down. The medical costs in the numerator of equation (7) must also be raised to be consistent with the revised denominator value. Since the estimate earlier was $0.7b consistent with 4,600 deaths, this implies $1.4b for 9,250 deaths.

In addition, GDP forecasts are required for March 2020 and from the same source just before the pandemic struck. Shortly before the pandemic arose, in December 2019, The Treasury (2019, page 3) forecasted New Zealand’s real GDP growth rates for 2020-2024 at the rates shown in the first row of Table 4. In April 2020, they released figures revised to reflect several different scenarios differing by the time for which Level 3 and 4 operated and the situation elsewhere in the world, of which the least severe (S1), the most severe (S3), and an intermediary scenario (S2) are shown in the other rows of the table (The Treasury, 2020, page 7). Arbitrarily designating 2019 GDP as 100, the GDP results under these paths are as shown in Table 4. The shortfalls between the 2019 forecasts and S1 are shown in the last row of the table and add to 27.8, which represents 28% of New Zealand 2019 GDP. Since New Zealand’s 2019 GDP was $311b, this is $87b.^27^ Even this figure is too low because these two forecast GDP paths have not fully converged by the end of the forecast period. All other scenarios considered by The Treasury yield even more severe GDP losses.

**Table 4:** GDP Forecasts

|  | 2020 | 2021 | 2022 | 2023 | 2024 | Sum |
| --- | --- | --- | --- | --- | --- | --- |
| Dec 2019 Forecasts | 2.2% | 2.8% | 2.7% | 2.5% | 2.4% |  |
| Implied GDP | 102.2 | 105.1 | 107.9 | 110.6 | 113.3 | 539.2 |
| April 2020 Forecasts: S1 | -4.5% | -2.5% | 10.0% | 5.5% | 4.0% |  |
| Implied GDP | 95.5 | 93.1 | 102.4 | 108.1 | 112.3 | 511.4 |
| April 2020 Forecasts: S2 | -8.0% | -3.0% | 13.0% | 5.5% | 4.0% |  |
| Implied GDP | 92.0 | 89.2 | 100.8 | 106.4 | 110.6 |  |
| April 2020 Forecasts: S3 | -8.0% | -23.5% | 30.0% | 13.0% | 6.5% |  |
| Implied GDP | 92.0 | 70.4 | 91.5 | 103.4 | 110.1 | 467.4 |
| Shortfall (2019 v S1) | 6.7 | 12.0 | 5.5 | 2.5 | 1.0 | 27.8 |

The proportion of this $87b loss arising from a lockdown rather than a mitigation policy must be estimated. The difference between the S1 and S3 scenarios shown in Table 4 is simply in how long Levels 3 and 4 persist (two months in S1 and twelve months in S3, which is a multiple of 6). In addition, going from S1 to S3 adds a further loss in GDP equal to 44% of the 2019 GDP (511.4 – 467.4), and this relates wholly to the last category of loss (the loss from lockdown rather than mitigation by the New Zealand government, with all other effects held constant). This suggests that the GDP loss within S1 arising from the New Zealand government’s adoption of a lockdown rather than a mitigation policy would be about 1/5 of that in moving from S1 to S3, which is 44%/5 = 8.8%. So, under S1, the GDP loss from lockdown rather than mitigation would be 8.8% of 2019 GDP, i.e., $311b*.088 = $27b. This represents 31% of the $87b GDP loss in S1, with the rest due to the other factors described earlier.

This estimate is conservative because the least severe scenario S1 has been used and its GDP forecast path has not fully converged on the December 2019 forecasts over the period for which the forecasts are available (out to 2024).

Substitution of all of these parameter values into equation (7) yields a cost per QALY saved of $660,000 as follows:

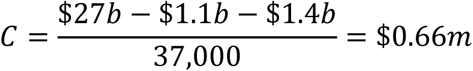

So, the cost per QALY saved is over 10 times the benchmark value of $62,000. This strongly favoured mitigation in March 2020. Allowing for the possibility that the lockdown would fail (Blakely et al, 2020, ascribed a 25% probability to this), the cost per QALY saved would be even higher and therefore mitigation would be even more strongly favoured.

This analysis characterizes the policy choice as lockdown or mitigation in March 2020. However there was an option to subsequently change policy, i.e., an initial lockdown could have been subsequently replaced by mitigation if the deaths (had mitigation been initially undertaken) were much lower than expected, and initial mitigation could have been subsequently replaced by lockdown if the deaths proved to be much larger than expected. I therefore modify the previous analysis, to allow for this possibility of changing the decision as information about the death toll unfolded.^28^ Blakely et al (2020) provide a range of possible deaths from mitigation, of at least 6,500, along with deaths of 500 if lockdown succeeded. The latter number of 500 is too small to materially affect the analysis, and is therefore set to zero, which is favourable to lockdowns. In respect of the range of deaths from mitigation if adopted throughout, I adopt the lower limit of 6,500 (because it does not materially affect the analysis) and designate the upper limit at *D_U_*. I also assume that the initial lockdown decision on 23 March is reconsidered after one month, at which point the death toll outcome from mitigation throughout (*D*) is then predictable, that switching from lockdown to mitigation at this point halves the GDP losses from the lockdown ($27b), that switching from lockdown to mitigation at this point does not lessen the death toll incurred from adopting mitigation (which is favourable to the decision to lockdown), that switching from mitigation to lockdown at this point does not reduce the GDP losses from the lockdown, and that switching from mitigation to lockdown at this point halves the death toll (which is generous to the lockdown switch decision in light of the evidence in the Appendix). The analysis appears in Table 5 below, with the first column showing the initial decision, the second column the deaths if mitigation were pursued throughout (*D*), the third column the decision after one month, the fourth column the cost from the policy sequence (deaths scaled up by four to QALYS and multiplied by the value per QALY of $62,000, plus GDP losses relative to mitigation throughout), and the fifth column the optimal choice in one month. Hospital costs, and GDP losses from work absences, are omitted because they are not material.

**Table 5:**
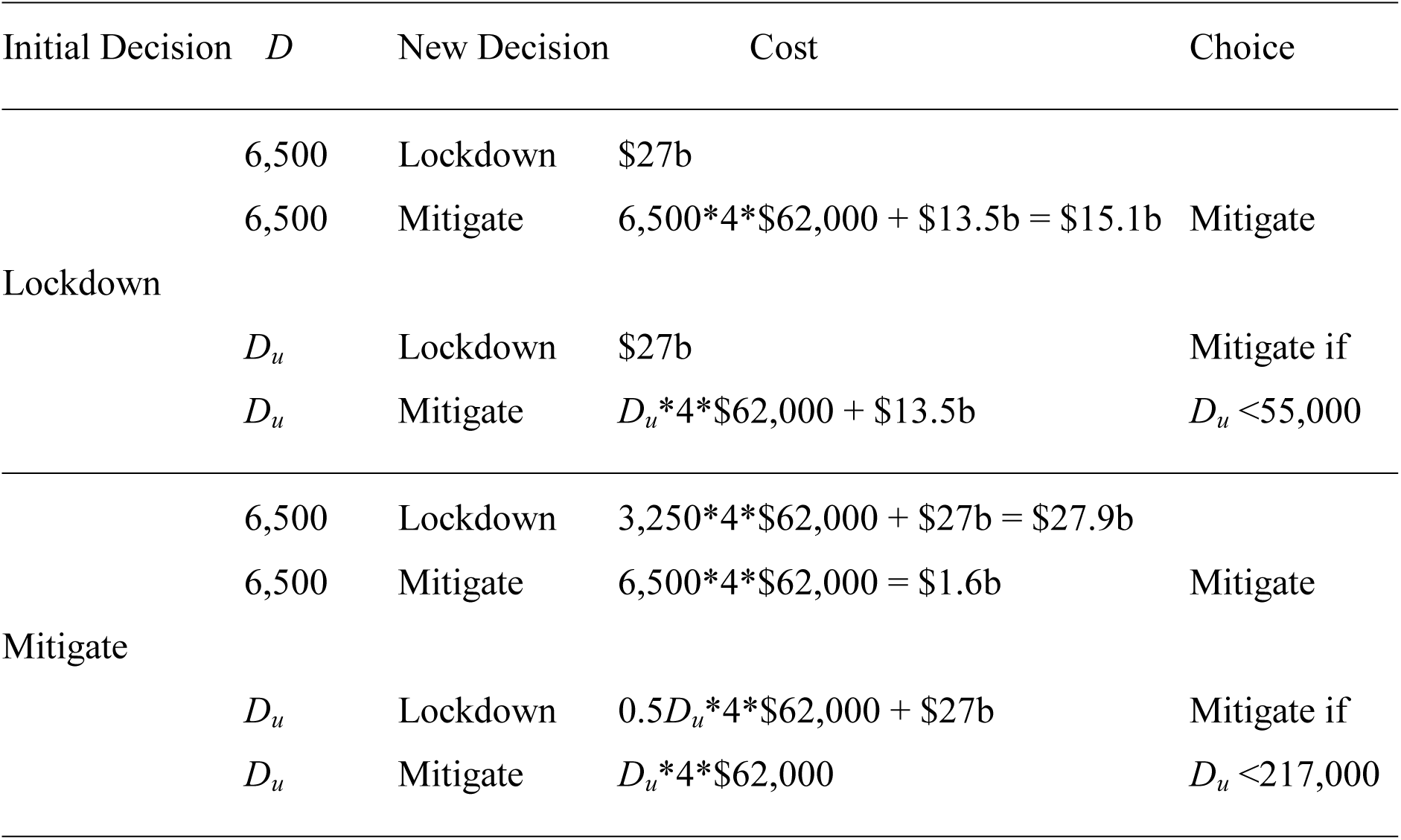
Optimal Decisions

As shown in the table, if *D_u_* < 55,000, the optimal decision in one month was always to mitigate, leading to costs if lockdown were initially adopted ranging from $15.1b (when *D* = 6,500) to $27b (if *D* = 55,000), and costs if mitigation were initially adopted ranging from $1.6b (when *D* = 6,500) to $13.6b (when *D* = 55,000). So, if *D_u_* < 55,000, the optimal initial decision would have been to mitigate, because its outcomes ($1.6b to $13.6b) were superior to their counterparts under lockdown initially ($15.1b to $27b), and mitigation would also have been optimal one month later. However, if 55,000 < *D_u_* ≤ 217,000, the optimal decision in one month if lockdown were initially undertaken would have been mitigation if *D* = 6,500 and lockdown if *D* = *D_u_*, yielding outcomes from locking down initially ranging from $15.1b to as much as $54b if *D* = *D_u_* = 217,000, whilst the optimal decision in one month if mitigation were initially adopted would have always been mitigation, yielding outcomes from mitigating initially that would have ranged from $1.6b to as much as $54b if *D* = *D_u_* = 217,000. Thus, if 55,000 < *D_u_* ≤ 217,000, mitigating initially would still have been superior, and it would have been followed by mitigating in one month. For lockdown initially to be optimal, *D_u_* would have to have exceeded 217,000 and substantially so to yield a probability distribution for *D* that favoured lockdown initially. This value for *D_u_* is well beyond any figure suggested by any expert observer in March 2020 (including the 83,000 under no controls in James et al, 2020, Table 2). Thus, there is no remotely plausible probability distribution for deaths under mitigation that would have justified lockdown initially. Blakely et al (2020) also alludes to the possibility of lockdown failing, but consideration of this possibility would have made lockdown initially even less favourable and therefore does not change the result here: mitigating initially was optimal, and remained so even if the decision could have been changed as death rate information unfolded, because the worst case death toll scenario was insufficiently high.

This leaves the question of why the New Zealand government chose a lockdown rather than a mitigation policy in March 2020. A natural candidate for explaining this is that it was extremely risk averse, i.e., it focused upon Blakely et al’s (2020) worst case death toll from mitigation over lockdown, of 12,500, which implies 50,000 QALYs saved by locking down. Substituting this into the denominator of the last equation, and scaling up the medical costs consistent with this incremental death toll of 12,500 (from $1.4b to $1.9b), the cost per QALY falls to $480,000:

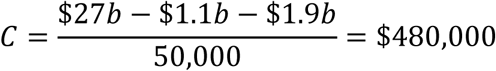

Even this is seven times the benchmark of $62,000. Even using Blakely et al’s (2020) highest figure of 30,100 deaths with no mitigating actions, implying 120,000 QALYs saved by locking down, and proportionately scaling up the health costs to $4.6b, the cost per QALY saved is $180,000, which is three times the benchmark. So, even extreme risk-aversion does not fully explain the government’s decision to lockdown.

A complementary possibility is that the New Zealand government was prepared to pay more than the worst-case scenario just described (at the conventional price of $62,000 per QALY saved) to buy ‘peace of mind’ for the whole population. Assuming that a lockdown would succeed, the additional payment would be *P* satisfying the following equation:

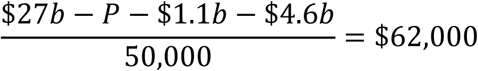

The solution is *P* = $18b, i.e., the peace of mind payment was $18b whilst the other $9b in GDP losses was payment to avoid the worst-case loss of 50,000 QALYs at $62,000 each. However, doing this is not consistent with standard methodology in assessing health interventions, and therefore with the ethical principle of equity. Furthermore, peace of mind benefits are unique to pandemics, as virtually all health or safety interventions increase peace of mind for the whole population as well as saving QALYs. For example, the road toll in New Zealand is currently about 370 per year, and peaked at 840 in 1973.^29^ This reduction has come in part through road safety expenditures, which have reduced deaths and therefore increased the peace of mind of every person (who is at risk of becoming a victim at any time they drive or even cross a road). Suppose such safety measures have saved 200 lives per year, with an average residual life expectancy for the victims of 40 years and perfect health, and the savings are accumulated over 50 years for comparison with pandemics (for which lockdowns might be contemplated) arising every 50 years. The QALY savings from the safety measures are then 200*40*50 = 400,000 QALYs over 50 years. This is vastly more than even the worst case mitigation scenario for Covid-19 of 50,000 QALYs saved. Despite this, road safety measures are evaluated purely on the basis of the value of the lives saved and not also because they increase peace of mind for the entire population.

In seeking an explanation for the decision to lockdown, it is noteworthy that the situation is characterized by several highly unusual features in health and safety interventions. Firstly, the source of the problem was new; the resulting absence of directly relevant historical experience may have led decision makers to fear worst case scenarios beyond even those articulated by expert opinion. Secondly, since the effectiveness of lockdowns could reasonably be expected to fall away quickly with any delay, a decision was required much more quickly than the speed at which governments normally operate and haste generally reduces the quality of decision making. Thirdly, the death toll from failure to lockdown would be more highly concentrated in time than most other health and safety interventions, which would exaggerate the emotional perception of the death toll and concentrate responsibility for it upon the decision makers, especially since the media gave Covid-19 deaths such attention. Fourthly, the cost of locking down (primarily in the form of GDP losses) would be distributed over several years and therefore would not be attributed entirely to the decision makers. Finally, the unusual form of the primary cost (GDP losses) lacks the emotional resonance from the more usual monetary payments, and therefore focused decision makers upon the benefits from lockdowns rather than costs as well as benefits. The confluence of these factors may have induced the decision to lockdown, contrary to standard principles of cost-benefit analysis even when using the worst case scenarios articulated by expert opinion.

## 4. Looking Forward on 28 June 2021

The analysis so far has examined the merits of lockdown versus mitigation being adopted by New Zealand in March 2020. Since then, there have been further but more localized lockdowns, and further instances are possible. Assessing the merits of any future lockdowns requires (as before) an estimate of the deaths that would be experienced under a mitigation policy. Such an estimate will be much less than that provided in the previous sections, even if the lockdown covered the same area and was for the same duration, for three reasons. Firstly, the period over which the virus could then inflict casualties would be much less, i.e., from the point in time at which it erupts (after 28 June 2021) until mass vaccination (of high-risk groups) is completed in several months, rather than from March 2020 until this mass vaccination point. Secondly, the vaccination campaign to date and to come over the period until mass vaccination is achieved will further reduce the death toll. Thirdly, the average death rate per day from now on would likely be less because policy and medical lessons have been learned since March 2020.^30^ In addition, although the GDP losses from such a lockdown will also be less than the lockdowns commencing in March 2020 ($17b), the reduction should be less pronounced than for the deaths because GDP losses are strongly tilted towards the lockdown period whilst the lives saved by lockdowns are more evenly spread over time until the mass vaccination point.

To illustrate these points, suppose that mass vaccination of high-risk groups will be completed at the end of 2021. Suppose further that a new outbreak that cannot be contained without a nation-wide lockdown occurs now, and adoption of a mitigation policy in response to it is expected to incur additional deaths (relative to a lockdown policy) equal to 28% of those incurred if a mitigation policy had been adopted in March 2020, because the period from now till the end of 2021 is 28% of the time period from March 2020 till the end of 2021. Since the additional deaths under the latter scenario (mitigation from March 2020) have been estimated at up to 4,600, the additional deaths under the former scenario (mitigation from the present time) are estimated at 28% of this, i.e., up to 1,300. The QALY losses from these additional deaths will be (as before) four times the number of deaths. In addition, suppose the medical costs for sufferers ($0.7b above) and GDP losses from those absent from work ($1.1b above) are scaled down in the same proportion as the deaths, to $0.2b and $0.3b respectively. Finally, suppose that GDP losses from this future lockdown will be 50% of the March 2020 lockdown, i.e., $8.5b. With no allowance for the effects of unemployment, the cost per QALY saved through a lockdown rather than mitigation follows equation (7) and would then be at least $1.54m as follows:

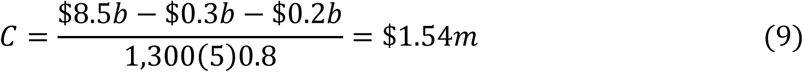

This is double the result in equation (7), and therefore favours a mitigation policy for future outbreaks even more strongly than in March 2020. If vaccinations along with policy and medical lessons learned since March 2020 further reduce the death toll, the denominator in (9) falls in proportion, raising the ratio in the same proportion, thereby favouring mitigation even more strongly.

By contrast, if an outbreak could be contained through locking down only part of the country, the GDP losses from doing so would be only some fraction (*P*) of the $8.5b in equation (9) whilst all other terms would be unchanged (because locking down any part of the country to prevent an outbreak that would otherwise spread to the entire country would warrant the same values for these other terms as for a nation-wide lockdown). The cost per QALY saved by lockdown would then be as follows:

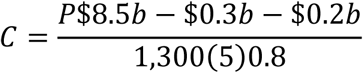

Thus, if *P* = 0.08, this cost would be $35,000, which is below the threshold of $62,000. So, lockdown would then be justified. In fact, lockdown would be justified for any value of *P* up to 0.10, i.e., a lockdown affecting a part of the country generating up to 10% of GDP would be warranted.

This analysis assumes that, if a lockdown occurs at some future point, only one such lockdown will be required before mass vaccination of the high-risk groups occurs. If more than one may be required, then the GDP loss of *P*$8.5b in the last equation would be increased. For example, if there were a 50% probability of a second lockdown affecting the same proportion of the country, the last equation would become

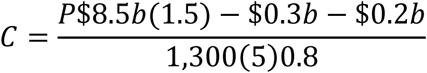

The lockdown policy would only then be justified if *P* were less than 0.07. Alternatively, if there were a 25% probability of a second lockdown affecting the entire country, the last equation would become

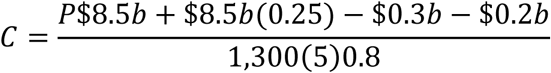

The lockdown policy would not then be justified for any value for *P*. All of this demonstrates that locking down only part of the country to contain an outbreak might seem justified, but much less so if one allows for the possibility of future outbreaks.

## 5. Conclusions

This paper has considered the costs and benefits of New Zealand’s lockdown strategy relative to pursuit of a mitigation strategy in March 2020. The estimated additional deaths from a mitigation policy are 1,750 - 4,600. The result is that the cost per Quality Adjusted Life Year saved by locking down is estimated to be at least 13 times the generally employed figure of $62,000 for health interventions in New Zealand; the lockdowns were therefore not justified. Consideration of the information available to the government in March 2020 yields a similar ratio and therefore strongly supported adoption of a mitigation strategy at that time. If New Zealand experiences a new outbreak, and cannot contain it without resort to a nation-wide lockdown, the death toll from adopting a mitigation strategy at this point would be even less than had it done so in March 2020, due to the vaccination campaign and because the period over which the virus would then inflict casualties would now be much less than the period from March 2020. This would favour a mitigation policy even more strongly than in March 2020. This approach of assessing the savings in quality adjusted life years and comparing them to a standard benchmark figure ensures that all quality adjusted life years saved by various health interventions are treated equally, which accords with the ethical principle of equity across people.

## Data Availability

All data referred to in the paper is publicly available at the weblinks provided in the paper.

https://www.worldometers.info/coronavirus/

https://covidtracker.bsg.ox.ac.uk/

https://www.euromomo.eu/graphs-and-maps#excess-mortality

## APPENDIX

This Appendix examines the possibility of reverse causality in equation (1), i.e., causality runs from *D* to *S* (as well as *S* to *D*), because

a. some governments chose their *S* value at the commencement of the crisis based upon their predictions of *D* under both low and high *S* scenarios, and/or
b. some governments chose their *S* values based upon their observation of their country’s death rate in the early stages of the crisis.

If either of these is true, the estimated coefficient on *S* in equation (1) may be biased. The traditional method of dealing with this is to use an “instrumental variable”, but no good candidates are apparent. I therefore enquire into the extent of these problems.

In respect of the first possible problem, I will focus upon the death rates under mitigation (*S* = 50) and extreme lockdown (*S* = 100). Suppose that there are two types of countries (A and B) whose governments held the views shown in Table 6 (at the commencement of the crisis) about expected death rates under mitigation and extreme lockdown.

**Table 6:**
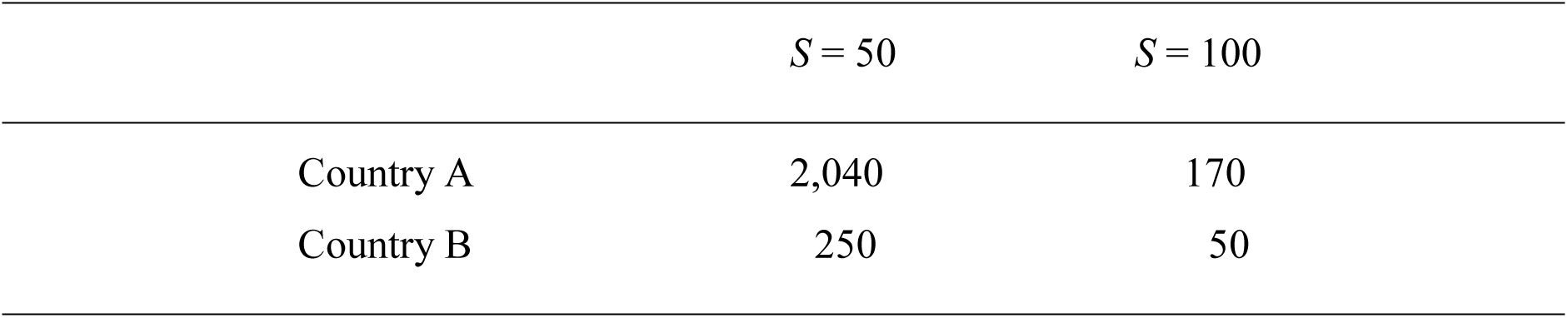
Expected Death Rates under Various Scenarios

Type A countries choose *S* = 100 because the expected death rate is unacceptably high with *S* = 50. Type B countries have much lower expected death rates than type A countries under both mitigation and extreme lockdown, and choose *S* = 50 because the expected death rate in that scenario is acceptable. If the governments’ predictions are on average accurate, then regressing *D* on *S* would then be expected to yield a coefficient on *S* of -1.6. However, the real coefficients on *S* are much lower, most particularly -37.0 for type A countries. Thus, the regression coefficient on *S* would be biased upwards.

It is plausible that some governments believed that their death rate under mitigation would be both large and vastly in excess of their death rate under extreme lockdown, as shown in Table 6, and acted accordingly in accordance with the predictions of experts like Ferguson et al (2020). It is also plausible that other governments believed that their death rates under mitigation would be much lower, as shown in Table 6, and acted accordingly in accordance with contrary expert opinions.^31^ However, both types of governments’ beliefs would need to be (on average) correct in order to be compatible with the expected coefficient on *S* in a regression like equation (1). Thus, there would have to be features of these two types of countries that would justify the markedly higher death rate in type A countries than in type B countries under mitigation (times 8 in Table 6), *and* governments would have to have been capable of recognizing these at the commencement of the crisis. Experts’ predictions, such as those of Ferguson et al (2020), would not have helped. For example, Ferguson et al (2020) used Chinese data to generate predictions for only the UK and US, which differed only slightly (3,700 per 1m for the UK and 3,500 for the US) due to demographics and population density (ibid, pp. 6-7 and 16). Furthermore, his numerous critics believed his death rates for the UK and US under mitigation were too high rather than that they were correct for those countries but far too high for others. Furthermore, if by some other means governments believed that their death rates under mitigation would markedly differ due to some variable other than the regressors used in equation (1) or those tested and rejected by me, and their beliefs were correct, they would have to have been aware in advance of a variable that I have not been able to locate even with the advantage of subsequently obtaining and testing the data that has become available since the commencement of the crisis. These conditions are not plausible, and this implies that the estimated coefficient on *S* in equation (1) is not materially biased for reasons of this type.

The second potential source of reverse causality in equation (1) is that some governments may have chosen their *S* values in light of their observation of their country’s death rate in the early stages of the crisis, believing it would predict the final death rate. To illustrate this, suppose there are two types of countries, with average death rates under mitigation and extreme lockdown as shown in Table 7. At the commencement of the crisis, it is unknown which category each country lies in but it is revealed by the death rates in the early stages of the crisis. So, upon observing their early stage death rates, the governments of type A countries then understood that they were of that type and chose extreme lockdown, yielding *S* = 100 and an average death rate of *D* = 170. At the same point, the governments of type B countries then understood that they were of that type and chose mitigation, yielding *S* = 50 and an average death rate of *D* = 250. Regressing *D* on *S* would then be expected to yield a coefficient on *S* of -1.6. However, the real coefficients on *S* are much lower, most particularly -8.6 for type A countries. Thus, the regression coefficient on *S* would be biased up.

**Table 7:** Average Death Rates under Various Scenarios

| | $S = 50$ | $S = 100$ |
| --- | --- | --- |
| Country A | 600 | 170 |
| Country B | 250 | 50 |

This scenario can be tested as follows. For each country, I regress its Stringency value ten days after its first reported death (*S_10_*) on its death rate up to that point (*D_10_*), to assess whether *D_10_* can explain *S_10_*. I repeat the process for 20 and 30 days after each country’s first death. I also test whether any of these three early stage death rates can explain the maximum *S* value chosen by governments (*S_m_*). These regressions yield the results shown in the first six columns of Table 8 below. Only two of these regressions even yield a positive coefficient on early death rate (consistent with the scenario in Table 8) and none of them yields a statistically significant coefficient on it. So, the hypothesis that early stage death rates did not affect governments’ choice of *S* cannot be rejected. This is surprising because the death rates up to day 20, and even more so for up to day 30, are good predictors of the death rate in the first wave of the crisis (to 22 August, and designated *D*), as shown in the last three columns of Table 8. So, at least from day 20, the death rate data up to that point could have been used to set the *S* value at that point or the maximum *S* value but governments did *not* seem to have done so.

**Table 8:** Stringency Levels and Death Rates

| Dep Var (DV) | $S_{10}$ | $S_{20}$ | $S_{30}$ | $S_m$ | $S_m$ | $S_m$ | $D$ | $D$ | $D$ |
| --- | --- | --- | --- | --- | --- | --- | --- | --- | --- |
| Indep Var (IV) | $D_{10}$ | $D_{20}$ | $D_{30}$ | $D_{10}$ | $D_{20}$ | $D_{30}$ | $D_{10}$ | $D_{20}$ | $D_{30}$ |
| Mean DV value | 68 | 77 | 79 | 81 | 81 | 81 | 198 | 198 | 198 |
| Mean IV value | 3 | 17 | 50 | 3 | 17 | 50 | 3 | 17 | 50 |
| Coeff on DV | 1.9 | -.07 | .002 | -.57 | -.10 | -.02 | 7.8 | 7.4 | 2.8 |
| $P$ Value for DV | .09 | .59 | .96 | .37 | .37 | .52 | .53 | 0 | 0 |
| Adjusted $R^2$ | .06 | -.02 | -.03 | -.01 | -.01 | -.02 | -.02 | .40 | .66 |

This raises the interesting question of what does then explain the maximum *S* values. Regressing this variable on the variables used in or tested for inclusion in equation (1), being population density, date of first death, proportion of population over 65, beds per 100,000 people, GDP per capita, and household size, yielded no statistically significant coefficients. However, ranking the maximum *S* values from highest to lowest reveals that the four countries arising from the breakup of Yugoslavia occupy four of the ‘top’ six slots (with an average *S* value of 95) and the five Scandinavian countries occupy four of the ‘bottom’ five slots (and have an average *S* value of 68). This suggests that the *S* choice was in part driven by mimicry of neighbouring countries. Consistent with this, Sebhatu et al (2020) finds that the speed with which restrictions were adopted by the OECD members was influenced by the behavior of nearby countries.

In summary, the cross-sectional regression in equations (1) does not seem to suffer from reverse causality from *D* to *S*.

## Footnotes

1 Within Europe, the mitigating countries have been Iceland, Finland, Latvia and Sweden, with death rates per 1m up to 28 June 2021 of 87, 175, 1,342 and 1,435 respectively. All death rate data in this paper are drawn from https://www.worldometers.info/coronavirus/.

2 See https://covidtracker.bsg.ox.ac.uk/ for the data. Their other indexes produce similar results.

3 Malta is excluded because Hale et al (2020a) does not include data on them. In addition the countries with very small populations (under 100,000) are excluded because many of the data sources used for this analysis do not provide data on them.

4 A closely related variable is the number of days from the date on which the Stringency Index reached 54 (the lowest of the cross-country maxima and therefore defined for all countries) until the date of the first death, because higher values indicate a faster response by a government to the crisis. Hale et al (2020b) use a similar variable. Each of these two variables is highly statistically significant and the *R*2 results from them are almost identical, but that from ‘Date of First Death’ are slightly better. I therefore report only the results from the use of ‘Date of First Death’.

5 For the population of countries, see the last column of https://www.worldometers.info/coronavirus/. For area of countries, see third column of https://en.wikipedia.org/wiki/List_of_countries_and_dependencies_by_area. For GDP per capita of countries, see the first column (IMF data) of https://en.wikipedia.org/wiki/List_of_countries_by_GDP_(nominal)_per_capita. For the population proportion over 65, see https://en.wikipedia.org/wiki/List_of_countries_by_age_structure. For average household size, see https://www.un.org/en/development/desa/population/publications/pdf/ageing/household_size_and_composition_around_the_world_2017_data_booklet.pdf<u>, pp. 20-24, except for Cyprus, which comes from</u> https://population.un.org/Household/index.html#/countries/196<u>. For the number of nursing and elderly home beds per 100,000 of population, see</u> https://gateway.euro.who.int/en/indicators/hfa_490-5100-nursing-and-elderly-home-beds-per-100-000/<u>, which does not contain data for Bosnia, Cyprus and Portugal so these numbers were estimated from those for Croatia, Greece and Spain respectively. For flu intensity, see Appendix to Hope (2020). The dates of the first deaths come from</u> https://www.worldometers.info/coronavirus/.

6 Using the average Stringency Index (from the first European death on 14 February to 30 December) instead of the maximum Stringency Index also yields a coefficient that is positive and statistically insignificant.

7 See https://www.euromomo.eu/graphs-and-maps#excess-mortality.

8 See https://www.ined.fr/en/everything_about_population/data/europe-developed-countries/population-births-deaths/.

9 Mass vaccination will at best reduce rather than eliminate deaths, because some people are unresponsive to vaccines or will not consent to them. However, it is very unlikely that lockdowns would be pursued beyond this point. So, in assessing the merits of lockdowns and mitigation, the relevant deaths are those up to this point.

10 See https://www.statista.com/statistics/1107913/number-of-coronavirus-deaths-in-sweden-by-age-groups/ and https://apps.who.int/gho/data/?theme=main&vid=61600. The age distribution is only available in ten-year blocks whilst life expectancy is only available in five year blocks up to age 85 followed by an 85+ group. So, Table 1 shows the number of victims in ten-year blocks up to age 80 as per the source data, the number of victims assigned to the 80-84 block is half of that reported in the 80-89 block, the other half of that block plus the 90+ block is combined to form an 85+ block, the life expectancies for the ten-year blocks up to age 80 are averaged over the data for each ten-year block, and the life expectancies for the last two blocks are as per the source data. The life expectancy data is also separately reported for males and females, unlike the age distribution of the victims, and the former data is therefore averaged over the sexes (since the Miles et al, 2020, data reveal that the sex split of the victims is close to 50/50, at 56% men).

11 The figure of 6.67 years in this column of the table is a weighted average of 0.6 years for the nursing home residents, with weight 6/21, and 9.1 years for the rest.

12 In respect of those dying in New York City up to 13 May 2020, and in those cases where the existing medical condition of the patient was known (no underlying condition or at least one underlying condition), 98% had at least one underlying condition (the set of conditions includes diabetes, cancer, heart disease, lung disease, and hypertension). See https://www.worldometers.info/coronavirus/coronavirus-age-sex-demographics/.

13 Data from the Period Life Tables 2012-2014, Table 5: http://archive.stats.govt.nz/browse_for_stats/health/life_expectancy/NZLifeTables_HOTP12-14/Data%20Quality.aspx#gsc.tab=0<u>. The table gives medians rather than means and therefore is not directly usable here.</u>

14 The figure of 4.72 years in the penultimate column of the table is a weighted average of 0.6 years for the nursing home residents, with weight 6/21, and 9.1(0.7) years for the rest. The figure of 2.80 in the final column is a weighted average of 0.6 years for the nursing home residents, with weight 13/21, and 9.1(0.7) years for the rest.

15 The figure for 2025 does not appear in the document, and is extrapolated from the series for comparison with the later forecasts, which do include 2025.

16 The GDP figure comes from Table M5 on the website of the RBNZ (www.rbnz.govt.nz).

17 The permanent displacement of other patients arises because the New Zealand public health system has insufficient capacity to even deal with the pre-covid demand, as is evident from the long queues. In respect of these queues, see https://www.nzherald.co.nz/nz/news/article.cfm?c_id=1&objectid=12365158.

18 See https://www.worldometers.info/coronavirus/.

19 The figure for 2025 does not appear in the 2019 document, and is extrapolated from the series for comparison with the forecasts one year later, which do include 2025.

20 For the Labour Force figure, see https://www.stats.govt.nz/information-releases/labour-market-statistics-disability-december-2020-quarter.

21 Frijters (2020) estimates that this loss reduces a person’s WELLBYs (a measure of happiness) by 0.7 and the loss of life by a healthy person would reduce WELLBYs by 6. So, the loss of employment for one year is equivalent to the loss of 0.7/6 = 0.12 years of healthy life.

22 This figure is used by The Treasury in its CBAx2020 model: see https://www.treasury.govt.nz/publications/guide/cbax-spreadsheet-model-0, tab “Impacts Database”, cell D135.

23 By comparison, Miles et al (2020, page 68) reports a guideline figure of 30,000 pounds used in hospitals in the UK (which is close to its 2019 GDP per capita of 32,000 pounds) and the larger figure of $125,000 in the US (ibid, page 72), which is approximately double its 2019 GDP per capita of $65,000.

24 See https://www.transport.govt.nz/mot-resources/road-safety-resources/road-deaths/, for 2020 to date.

25 See Tables 5 and 6 of the NZ Periodic Life Tables: http://archive.stats.govt.nz/browse_for_stats/health/life_expectancy/NZLifeTables_HOTP12-14.aspx#gsc.tab=0.

26 Pharmac (2015, pp. 51-52) recommends that all costs and benefits in health expenditure assessments be discounted by 3.5% per year, based on the five-year average real government bond rate. The longest term inflation-indexed bonds for which data is available for the last five years are those maturing in September 2035, and the average rate over the last five years has been 1.58% (see Table B2 on the website of the RBNZ: www.rbnz.govt.nz).

27 The GDP figure comes from Table M5 on the website of the RBNZ (www.rbnz.govt.nz).

28 Davies and Grimes (2020) also analyse this scenario, but their analysis is generic rather than specific to the situation prevailing in March 2020.

29 See https://www.transport.govt.nz/statistics-and-insights/safety-road-deaths/death-on-nz-roads-since-1921/.

30 Examples include the importance of properly quarantining rest homes, the best use of ventilators, and the merits of awake prone positioning. See https://www.statnews.com/2020/04/08/doctors-say-ventilators-overused-for-covid-19/<u>, and</u> https://www.nzherald.co.nz/nz/news/article.cfm?c_id=1&objectid=12322501<u>, and Koeckerling et al (2020).</u>

31 See for example https://thehill.com/opinion/healthcare/489962-what-if-the-sky-is-falling-coronavirus-models-are-simply-wrong.

